# A novel echo-parameter supersedes the Society of Thoracic Surgeons risk score in predicting post transcatheter aortic valve replacement mortality

**DOI:** 10.1101/2021.09.24.21264084

**Authors:** Chieh-Ju Chao, Pradyumma Agasthi, Amith R Seri, Timothy Barry, Anusha Shanbhag, Yuxiang Wang, Mackram Eleid, David Fortuin, John P. Sweeney, Peter Pollak, Abdallah El Sabbagh, Steven J. Lester, William K Freeman, Tasneem Z. Naqvi, David R Holmes, Christopher P. Appleton, Reza Arsanjani

**Affiliations:** Department of Cardiovascular Diseases, Mayo Clinic Arizona, Scottsdale, Arizona; Department of Cardiovascular Diseases, Mayo Clinic Rochester, Rochester, Minnesota; Department of Cardiovascular Diseases, Mayo Clinic Florida, Jacksonville, Florida

**Keywords:** Aortic valve stenosis, transcatheter aortic valve replacement (TAVR), STS risk score, augmented mean arterial pressure, mortality

## Abstract

**Objective:** To accurately predict post-TAVR mortality, we proposed a family of new echo-parameters (augmented blood pressure) derived from blood pressure and aortic valve gradient measurements and examined them in this study.

**Patients and Methods:** Patients in the Mayo Clinic National Cardiovascular Diseases Registry-TAVR database who underwent TAVR between January 1, 2012, and June 30, 2017, were identified to retrieve baseline demographics, echocardiographic and mortality data. Augmented blood pressure parameters and valvulo-arterial impedance were evaluated by univariate and multivariate Cox regression. Receiver operating characteristic curve analysis was used to assess the model performance against the Society of Thoracic Surgeons (STS) risk score.

**Results:** The final cohort contained 974 patients with a mean age of 81.4±8.3 years old, and 56.6% were male. The mean STS risk score was 8.2±5.2. The median follow-up duration was 354 days, and the one-year all-cause mortality rate was 14.2%. Both univariate and multivariate Cox regression showed that augmented systolic blood pressure and augmented mean arterial pressure (AugMAP) parameters were independent predictors for 1-year post-TAVR mortality (all p<0.0001). A univariate model of AugMAP1 supersedes the STS score model in predicting 1-year post-TAVR mortality (area under curve: 0.700 vs. 0.587, p=0.0051).

**Conclusion:** Augmented mean arterial pressure provides a simple but effective approach for clinicians to quickly estimate the clinical outcome of TAVR patients. It can be incorporated in the assessment of TAVR candidacy.

## Background/Introduction

The success of transcatheter aortic valve replacement (TAVR) has substantially changed the landscape of management of aortic valve disease^1–3^. With the expansion of TAVR indications, it is anticipated that more TAVR procedures will be performed in the foreseeable future ^4^. Given the underlying comorbidities, the clinical outcomes after the TAVR procedure have gained significant attention that involves both conventional and machine learning research approaches ^5– 12^. While the Society of Thoracic Surgeons (STS) risk score was initially designed to risk-stratify patients for surgical aortic valve replacement, the score was reported to be a strong predictor for short and long-term post-TAVR prognosis ^13–15^. In addition, many studies have reported valvulo-arterial impedance (Zva) as a predictor for post-TAVR prognosis ^8–10,16^. High Zva is associated with worse quality of life and exercise performance at one-year post-TAVR ^10^, while there are inconsistent results in predicting long-term mortality ^17–19^. Our group previously reported that cardiac power index and gradient-adjusted cardiac power index are good predictors of 1-year mortality after TAVR ^6,7^. In calculating gradient-adjusted cardiac power index, transvalvular gradient (mean transvalvular gradient or instantaneous peak transvalvular gradient) was added to the systolic blood pressure as augmented systolic blood pressure calculate augmented mean arterial pressure (AugMAP). The augmented MAP component is conceptually close to the summation of valvular and arterial load, which is the numerator of the Zva formula. Nagura et al. suggested that Zva is sensitive to the change of stroke volume index but not the arterial load. The potential measuring error from stroke volume index can be magnified in Zva calculation, especially in patients with low-flow status ^18^. In this context, we aimed to investigate the prognostic value of augmented blood pressure in TAVR patients and compare it to Zva and the STS risk score. We hypothesized that augmented systolic blood pressure and augmented mean arterial pressure can accurately predict the 1-year mortality in patients who underwent a TAVR procedure.

## Materials and Methods

### Study population, baseline demographics, and clinical data

A chart review was conducted on patients included in the Mayo Clinic National Cardiovascular Diseases Registry (NCDR)-TAVR database, which included patients from three major academic medical centers located in Rochester, MN, Phoenix, AZ, and Jacksonville, FL. We identified all patients aged ≥18 years who underwent TAVR between January 1, 2012, and June 30, 2017. Baseline demographics, lab data, device data, STS risk score, and follow-up data were directly extracted from the database. Patients who had prior TAVR procedure(s) were excluded. The Institutional Review Board at Mayo Clinic approved the study protocol, and all the patients provided research authorization to utilize their medical information.

### Baseline transthoracic echocardiography

Baseline transthoracic echocardiography (TTE) with 2-dimension imaging and Doppler were performed pre-procedure using commercially available ultra-sound scanners (Philips iE33; Phillips Medical Systems, Andover, MA, USA; GE Vivid E9, GE Healthcare, Milwaukee, WI, USA). All echocardiograms were interpreted according to the guidelines ^20–22^. Offline measurements of the images were obtained using ProSolv Cardiovascular Analyzer 3.0 (ProSolv Cardiovascular Inc., Indianapolis, IN, USA).

### Calculation of augmented systolic blood pressure, mean arterial pressure, and valvulo-arterial impedance

Non-invasive blood pressure measured at the time of baseline TTE was used to calculate augmented blood pressure parameters. The augmented blood pressure calculation formulas are stated as below:

1. Augmented SBP1(AugSBP1): Mean aortic valve gradient (mean AVG) was added to systolic blood pressure **(Equation 1)** and augmented MAP1(AugMAP1) was calculated by replacing the SBP with augmented SBP1 in the MAP formula **(Equation 3);**
2. Augmented SBP2(AugSBP2): Aortic valve maximal instantaneous gradient was added to systolic blood pressure (**Equation 2**), and augmented MAP2(AugMAP2) was calculated by replacing the SBP with augmented SBP2(**Equation 4**); and
3. Augmented MAP3(AugMAP3): Aortic valve mean gradient was added to mean arterial pressure (**Equation 5**) ^6^. Zva was calculated according to the standard formula by dividing the sum of the systolic blood pressure and mean transvalvular gradient by stroke volume index (SVi)^18^.

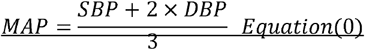

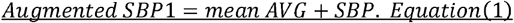

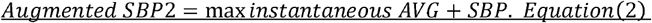

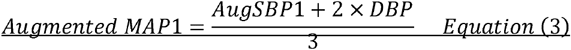

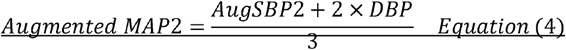

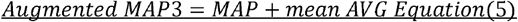

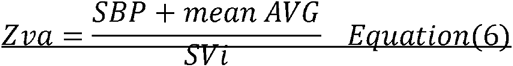

### Statistical analysis

Patients were grouped into alive and deceased groups and analyzed accordingly. All the two-group comparisons were summarized as alive group versus deceased group if not otherwise specified. Continuous variables were summarized as mean ± standard deviation, and the differences among groups were evaluated with Student t-test and Mann-Whitney U test for normally and non-normally distributed data, respectively. Categorical variables were expressed as counts and percentages, and differences among groups were evaluated with the chi-square test. Kaplan-Meier survival curve and Cox regression were used for survival analysis; the median of AugSBP1 and AugMAP1 were used as the cutoffs to stratify the patients for Kaplan-Meier analysis. In multivariate Cox regression analysis, variables were adjusted for age, sex, and STS risk score (Python lifelines 0.26.3). The augmented blood pressure measurements, Zva, and the STS risk score were used to develop univariate regression models separately. Receiver operating characteristics (ROC) curve analysis with the area under the curve (AUC) was used to assess the accuracy of each univariate Cox regression model against the STS risk score model. The bootstrap method was used to estimate the 95% confidence interval of AUC for each univariate model. DeLong’s test was used to assess the difference in model performance against the STS risk score model. A p-value of less than 0.05 was used as the cutoff of statistical significance for all the hypotheses. All the analyses were performed in Python version 3.7.10.

## Results

### Study Population and Baseline Demographics

A total of 974 patients were included for the final analysis after excluding 97 patients who had prior TAVR procedure(s). The mean age was 81.4±8.3 years old, 56.6% were male (n=551), and 97.3% (n= 947) were white. The mean STS risk score was 8.2±5.2. The median follow-up duration was 354 days (interquartile range 51-378 days), and the one-year all-cause mortality rate was 14.2% (n=139). The median time from baseline TTE to TAVR was 1.46 months. Systolic blood pressure (alive versus deceased: 130.9±21.4 mmHg vs. 119.1±20.0 mmHg, p<0.0001), diastolic blood pressure (68.9±13.1 mmHg vs. 62.3±12.3 mmHg, p<0.0001), and mean arterial pressure (89.5±13.4 mmHg vs. 81.2±13.0 mmHg, p<0.0001) were significantly higher in the alive group. Detailed data are summarized in **Table 1**.

### Augmented blood pressure, echocardiography, and Zva measurements

There was no significant difference in LV ejection fraction (alive versus deceased: 57.8±12.8% vs. 55.9±14.5%, p= 0.2214), mean aortic valve gradient (44.2±13.1 mmHg vs. 42.7±12.6 mmHg, p= 0.1988), maximal aortic valve instantaneous gradient (72.1±21.1 mmHg vs. 69.1±21.0 mmHg, p=0.0803), and stroke volume index (43.7±9.8 ml/m^2^ vs. 42.8±9.7 ml/m^2^, p= 0.1232). Both right atrial pressure (7.2±3.8 mmHg vs. 8.9±5.2 mmHg, p=0.0004) and right ventricular systolic pressure (41.0±13.2 mmHg vs. 45.4±18.5 mmHg, p=0.0097) were significantly higher in the alive group.

Regarding augmented blood pressure measurements, both AugSBP1 (174.9±25.6 mmHg vs. 161.5±23.9 mmHg, p<0.0001) and AugSBP2 (203.0±30.4 mmHg vs. 187.2±30.0 mmHg, p<0.0001) were significantly higher in the alive group. There were similar findings for AugMAP1 (104.1±14.3 mmHg vs. 95.3±13.7, p<0.0001), AugMAP2 (113.3±15.3 mmHg vs. 103.8±15.1 mmHg, p<0.0001) and AugMAP3 (133.5±19.3 vs. 123.6±18.1, p<0.0001). Box plots are used to visualize the augmented blood pressure data (**Figure 1**). Higher Zva was also observed in the alive group (4.2± 1.1 mmHg ml^-1^ m^-2^ vs. 3.9± 0.9, p=0.0233).

**Figure 1.**
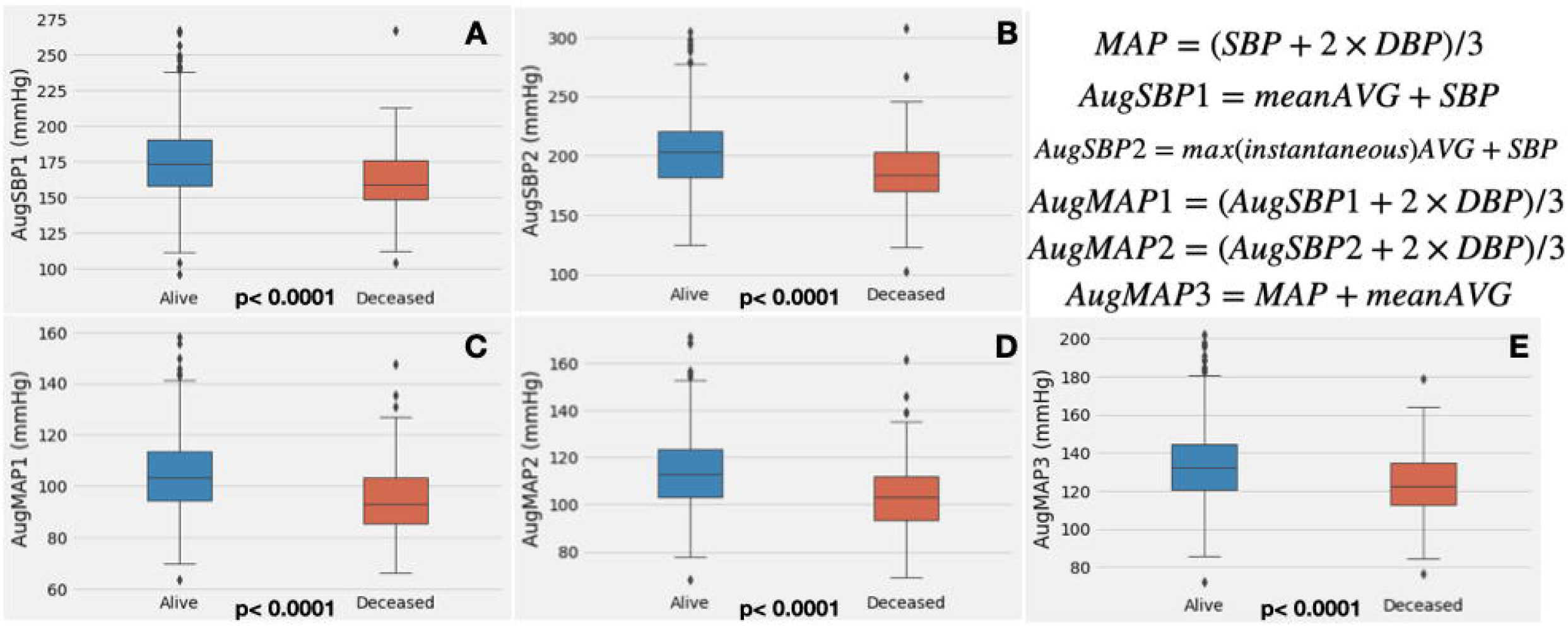
Box plot of augmented blood pressure measurements. Panel A-E demonstrates the box plot of each augmented blood pressure parameter. Augmented blood pressure parameters were significantly higher in alive patients when compared to deceased patients (all p<0.0001). The formulas used to calculate each parameter are listed at the right upper corner.

### Kaplan-Meier analysis and Cox regression

The median of AugSBP1 was 171 mmHg, and the median of AugMAP1 is 102.5 mmHg. In Kaplan-Meier analysis, both parameters demonstrated significant survival differences when comparing patients with ≥ cutoff vs. patients < cutoff (all p<0.0001) (**Figure 2A** and **2B**). In univariate Cox regression, AugSBP1, AugSBP2, AugMAP1, AugMAP2, and AugMAP3 were independently associated with 1-year all-cause mortality. These associations remained significant after adjusting for age, sex, and STS risk score in multivariate Cox regression (all p< 0.0001). In contrast, while Zva was independently associated with 1-year mortality in univariate regression (HR 0.82, 95%CI: 0.67-1.00, p=0.0456), the significance was not remained in multivariate Cox regression (HR 0.83, 95%CI: 0.68-1.00, p=0.0536). Stroke volume index was not independently associated with 1-year mortality in univariate Cox regression (HR 0.99, 95%CI: 0.97-1.01, p=0.3475). **Table 2** summarizes the Cox regression data.

**Figure 2.**
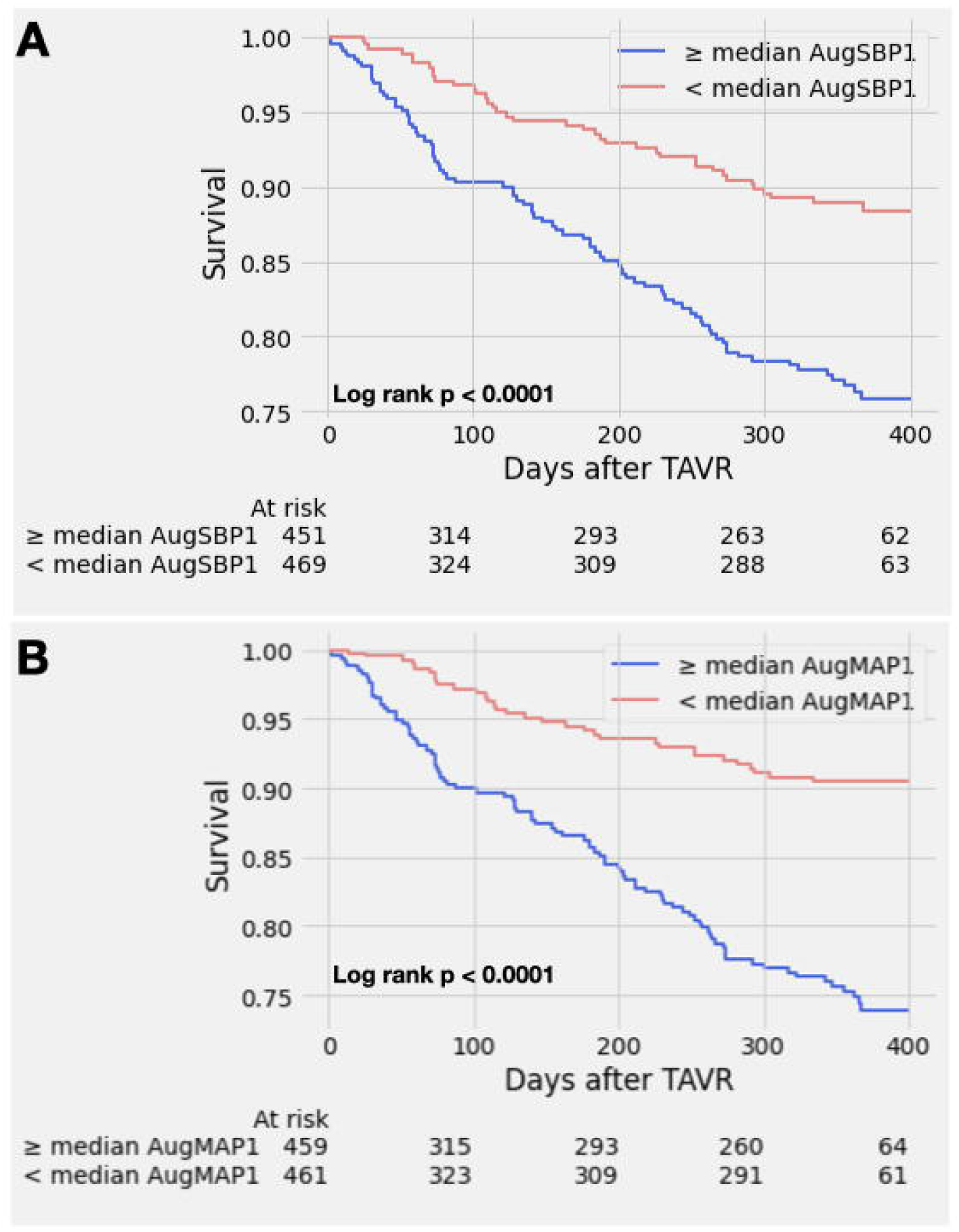
Kaplan-Meier survival curve analysis. **Panel A**. demonstrates significant survival difference between patients with ≥ median AugSBP1 vs. patients with < median AugSBP1; the median of AugSBP1 is 171 mmHg (Log rank p< 0.0001). **Panel B**. demonstrates significant survival difference between patients with ≥ median AugMAP1 vs. patients with < median AugMAP1; the median of AugMAP1 is 102.5 mmHg (Log rank p< 0.0001)

### Performance of univariate Cox regression models

The STS score univariate Cox regression model (reference model) had an AUC of 0.587 (95%CI 0.521 - 0.649) in predicting 1-year mortality. Among all other univariate Cox regression models, the AugMAP1 model (AUC 0.700, 95%CI 0.646 - 0.750, p=0.0051) and AugMAP2 model (AUC 0.691, 95%CI 0.636 - 0.743, p=0.0094) significantly outperformed the STS score model. AugSBP1 (AUC 0.665, 95%CI 0.612 - 0.719, p=0.052), AugSBP2 (AUC 0.649, 95%CI 0.599 - 0.704, p=0.1182) and AugMAP3 (AUC 0.654, 95%CI 0.597 - 0.709, p=0.086) were comparable/non-inferior in predicting mortality when compared to the STS score model. The AUC for Zva (0.559, 95%CI 0.502 - 0.611, p=0.519) was numerically smaller than the STS score; however, the difference was not statistically significant. **Figure 3**. demonstrates all the ROC curves.

**Figure 3.**
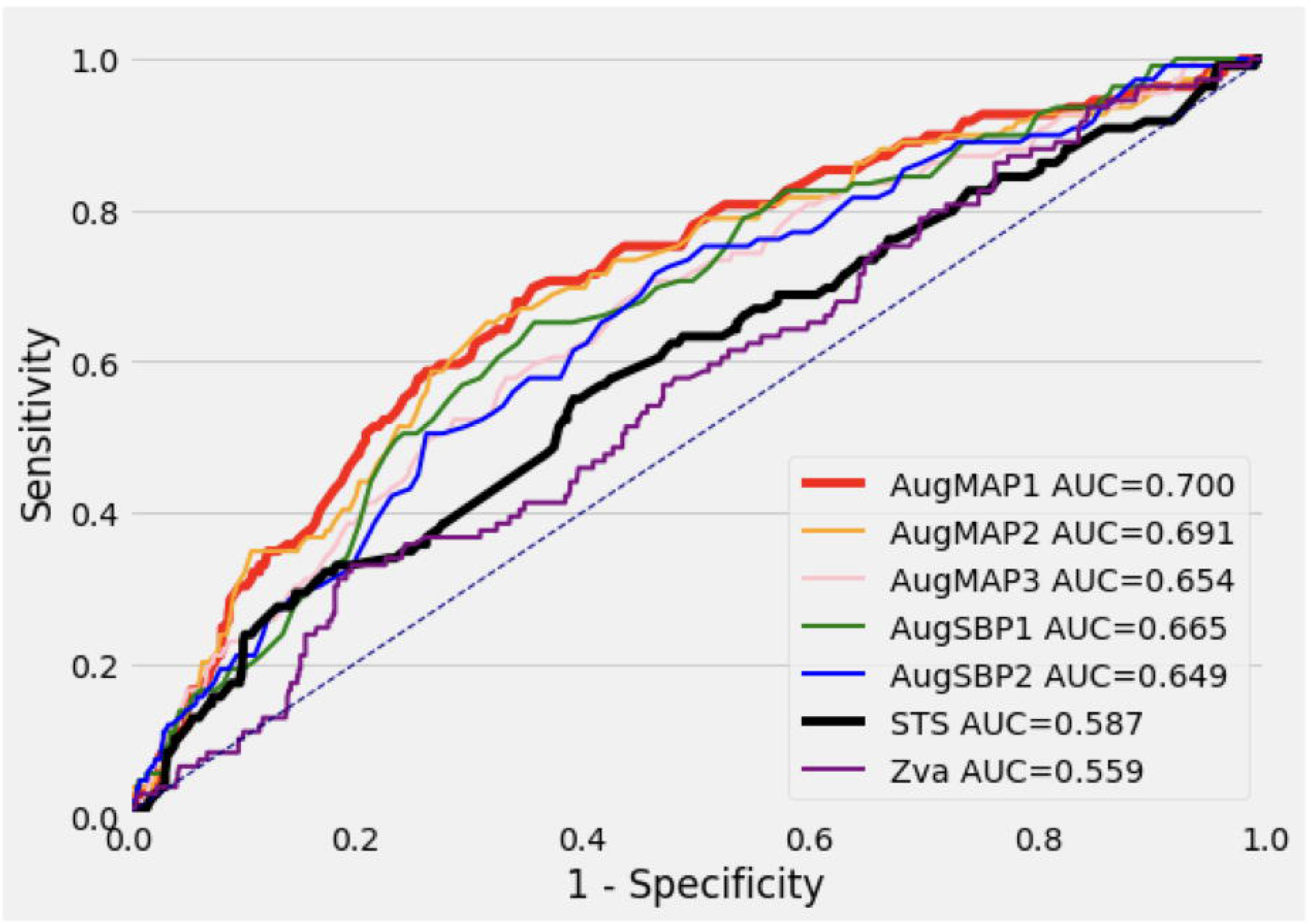
The ROC curves of all the single-parameter prediction models against STS risk score model. The ROC curves of all the single-parameter prediction models against the STS risk score model (AUC 0.587, 95%CI 0.521 - 0.649). The AugMAP1 model had the best performance (AUC 0.700, 95%CI 0.646 - 0.750, p=0.0051), followed by the AugMAP2 model (AUC 0.691, 95%CI 0.636 - 0.743, p=0.0094). Rest of the augmented blood pressure (AugMAP3, AugSBP1, AugSBP2) parameters were comparable to the performance of STS risk score (larger AUC, no statistical significance). Valvulo-arterial impedance (Zva) had smaller AUC than STS risk score, but this was not statistically significant (AUC 0.559, 95%CI 0.502 - 0.611, p=0.519).

## Discussion

In this retrospective study, we demonstrated that baseline augmented mean arterial pressure (AugMAP1) superseded the STS risk score in predicting 1-year post-TAVR all-cause mortality. To the best of our knowledge, this is the first application of augmented blood pressure parameters in assessing post-TAVR mortality. Our findings suggest that AugMAP1, as a surrogate marker of cardiac contractile function against systemic afterload in aortic stenosis patients, is closely related to post-TAVR mortality. Thus, AugMAP provides a simple but effective approach, which can be potentially incorporated in assessing TAVR candidacy.

### The physiology meaning of augmented blood pressure

When calculating the augmented blood pressure, we assumed that adding either mean or maximal instantaneous gradient to the systemic systolic blood pressure can reflect the true systolic pressure generated by the left ventricle ^6^. While non-invasive MAP is practically calculated as the summation of diastolic blood pressure and 1/3 pulse pressure, MAP also largely equals the product of cardiac output and systemic vascular resistance (SVR) (assuming a relatively small central venous pressure level). Therefore, MAP can be considered the capability of the left ventricle to generate cardiac output based on a given systemic vascular resistance, as MAP is proportional to cardiac output when systemic vascular resistance is constant. In the setting of aortic stenosis, a higher augmented MAP indicates the better cardiac contractile function to generate higher blood pressure against the afterload (stenotic aortic valve and SVR), and augmented MAP is substantially the true mean pressure of the left ventricle in a cardiac cycle. Therefore, among the three AugMAP parameters, AugMAP1 should be considered the most accurate gauge of left ventricular contractile function against the systemic afterload and thus most closely associated with the post-TAVR prognosis. The decremental model performance (AUC) of the univariate Cox models of AugMAP 1, 2, and 3 reflected that any deviation (change of the formula) from AugMAP1 would lead to weaker associations between the parameter and post-TAVR mortality. Also, for AugSBP1 and AugSBP2, while the AUC of each model was still larger than the STS model, the difference did not reach statistical significance as AugMAP1 and AugMAP2. This can be explained by the same theory. Since the diastolic blood pressure component was removed from the formula, the AugSBP measurements were deviated to the end of systolic blood pressure and did not truly reflect the real MAP/contractile function.

### An overlooked outcome predictor: augmented blood pressure

In both univariate and multivariate Cox regression analysis, all the augmented blood pressure parameters were shown to be independent predictors of 1-year post-TAVR mortality, and AugMAP1 had the best performance among them. Importantly, AugMAP1 also superseded the STS risk score in predicting 1-year mortality in head-to-head ROC curve comparison (p=0.0051). As the standard approach of TAVR candidacy assessment, the STS score is also reported to be a strong predictor for both short-and long-term post TAVR mortality^13–15^; therefore, it was selected as the reference model in this study. Hemmann et al. reported that the STS score model had an AUC of 0.679 (95% CI: 0.610-0.748) in predicting 1-year post TAVR mortality based on a cohort with 426 patients and with a performance superior to the EuroSCORE and EuroSCORE2^14^. In another study with 3491 TAVR patients, the STS score model had an AUC of 0.61 (95% CI: 0.56-0.67) in predicting 30-day post-TAVR mortality ^15^. Overall, the AUC of the STS score models in prior studies was in the range of 0.6-0.7, similar to the finding in our study (AUC 0.587, 95%CI 0.521 - 0.649). Furthermore, the performance of the univariate AugMAP1 model was not inferior to any STS score models above ^13–15^ and almost reached the same level as our previously published machine learning model developed from the same database ^5^. Using the median cutoff values of AugSBP1 (171 mmHg) and AugMAP1 (102.5 mmHg), we were able to stratify the post-TAVR prognosis in Kaplan-Meier survival analysis (**Figure 1A** and **1B**). The two parameters are simple, easy to calculate, and surprisingly accurate, with only a few variables as their input.

A possible concern for this approach is that the parameter primarily relies on blood pressure measurements, which can vary from time to time, and suboptimal blood pressure control may cause higher blood pressure readings. In our cohort, all of SBP, DBP, and MAP measurements were significantly higher in the alive group (all p< 0.0001), however the mean SBP in the alive group was reasonably well controlled (130.9±21.4 mmHg). There was no significant difference between the alive and deceased groups regarding antihypertensive medications. However, considering augmented blood pressure as the surrogate marker of cardiac contractile function, we would argue that higher (but reasonably controlled) baseline blood pressure stands for better cardiac contractile function in these patients and can potentially be related to a better outcome. This concept is supported by earlier studies that patients with higher blood pressure/hypertension after TAVR procedure was associated with better outcomes^23^, in contrast to lower blood pressure^9^. Furthermore, patients who developed hypertension post TAVR procedure were found to have significantly improved cardiac output and stroke volume^23^, which suggests these patients had better cardiac contractile function reserved, and therefore were able to generate higher blood pressure after relieving stenosis at the level of valve, in contrast to those with lower blood pressure^9^.

### Valvulo-arterial impedance (Zva) vs. AugMAP/AugSBP

In 2009, the concept of Zva was introduced by Hachicha et al., and this parameter was shown to be independently associated with mortality in aortic stenosis patients who underwent either surgical aortic valve replacement or medical therapy^24^. After that, Zva was evaluated in multiple studies as a post-TAVR outcome predictor ^10,17–19,25,26^. A cohort study containing 202 TAVR patients reported that patients who died within 6 months had higher baseline Zva and less improvement in post-TAVR Zva. While the authors reported that baseline Zva was independently associated with 6-months all-cause mortality, this conclusion was only based on univariate logistic regression and was not adjusted for other covariates^19^. In terms of longer-term mortality, Katsanos et al. showed that higher baseline Zva was independently associated with mortality at 2 years^17^ However, their study also had a smaller patient cohort (n=116), and the mortality endpoint only happened in 21 (18%) patients. Another study based on the Optimized CathEter vAlvular iNtervention (OCEAN)-TAVI registry included 1004 patients but reported that post-TAVR Zva was not associated with two-year all-cause mortality in multivariate analysis; the correlation between baseline Zva and mortality was not assessed in this study ^18^ Importantly, none of the above studies had used Zva to generate a single-parameter model and compare the performance with the STS risk score. In the current study ^17–19^, we observed that baseline Zva was an independent predictor of 1-year post-TAVR mortality in univariate Cox regression, but not after adjusting for age, sex, and the STS score. Furthermore, the univariate Zva model had the worst performance, while not statistically significant compared to the STS model (0.559 vs. 0.587, p=0.519). Compared to the performance of augmented blood pressure measurements, the component of total load (SBP + mean AVG, equals to AugSBP1) is likely the key portion for Zva to be associated with mortality in prior works^17–19^. Introducing stroke volume index as the denominator substantially canceled the contribution of cardiac output from the total load term and can bring additional measuring error to this parameter, especially in patients with low-flow status ^18^. Our true MAP-prognosis theory anticipates that the best accuracy Zva could achieve is at the level of AugSBP1 (the numerator of the Zva formula). However, with the contribution and potential measurement error from the stroke volume index, Zva deviates more from the augmented MAP thus had worse performance than AugSBP1. Our results comparing AugMAP and Zva also support the concept that cardiac contractile function against the afterload is more important than the vascular load in determining the outcome of TAVR patients.

### Incorporating augmented MAP in the assessment of TAVR patients

Compared to the STS risk score, which requires an input of more than 70 variables and relies on an online calculator after completing extensive workup^27^, our models with augmented MAP and augmented SBP provide a simple and effective way that only requires 2-3 readily available variables (SBP, DBP and mean AVG) to make a superior prediction. The median cutoffs (AugSBP1: 171 mmHg and AugMAP1: 102.5 mmHg) can be easily calculated at the bedside to provide real-time assessment, which may facilitate workup and decision-making, and can potentially change the assessment of TAVR candidacy.

## Conclusion

As a single parameter, augmented mean arterial pressure (AugMAP1) supersedes the STS risk score in predicting 1-year post-TAVR mortality. Augmented blood pressure parameters provide a simple but effective approach for clinicians to quickly estimate the clinical outcome of TAVR patients and can be potentially incorporated in the assessment of TAVR candidacy.

## Limitations

This study is limited by its retrospective nature, and external validation is not available for the findings. To maintain a sufficient sample size, we did not further divide our patient cohort into subgroups for ROC analysis. Concerning the period of TAVR procedures, most of the patients in this cohort were considered intermediate-to-high risk, so low-risk patients may not be well represented in this development cohort. The time from the baseline TTE/ blood pressure measurement to the TAVR procedure varied among patients, and the potential changes in between were not considered. However, the median time from baseline TTE to TAVR was relatively short (1.46 months) for significant hemodynamic changes. While our approach eliminated the contribution of stroke volume index, the component of systolic blood pressure and transvalvular gradient in this parameter still inherited the intrinsic limitation of Zva^28^. Also, invasive hemodynamic measurements were not available to validate the correlations between non-invasively measured augmented MAP and the invasive MAP, which is a potential direction for future studies. In addition, only all-cause mortality was available as an endpoint; whether augmented blood pressure parameters were related to other endpoints (cardiovascular mortality, heart failure hospitalization, etc.) shall be deferred to future studies.

## Data Availability

NA

## Abbreviations

AVG: aortic valve gradient
AugSBP: augmented systolic blood pressure
AugMAP: augmented mean arterial pressure
CO: cardiac output
DBP: diastolic blood pressure
MAP: mean arterial pressure
SBP: systolic blood pressure
SVi: stroke volume index
STS: Society of Thoracic Surgeons
SAVR: surgical aortic valve replacement
TTE: transthoracic echocardiography
TAVR: transcatheter aortic valve replacement
Zva: valvulo-arterial impedance

